# Disentangling the contribution of disease genes to drug therapeutic and side effects

**DOI:** 10.64898/2026.05.04.26352378

**Authors:** Panagiotis N. Lalagkas, Rachel D. Melamed

**Author notes:** Corresponding author: Rachel D. Melamed.

## Abstract

Most clinical trials fail due to either lack of efficacy or safety concerns. Human genetics can address both failure reasons: disease-associated genes are not only promising therapeutic targets but also predict drug side effects. However, because the same genetic signal underlies both outcomes, we need methods that disentangle which disease genes mediate therapeutic benefit versus adverse side effects. We use DraphNet, our previously developed model that maps drug molecular effects onto disease genes to generate two gene sets per drug: one linked to its therapeutic effects (IND genes) and one linked to its side effects (SE genes).

We show that IND and SE genes overlap for 76% of the tested drugs (compared to a null model). We also show that drugs sharing greater IND similarity also have greater SE similarity (ρ=0.57, p<1e-300). To show how our approach enables insights into drug biology, we construct groupings of drugs based on their IND and SE genes. We find that drugs in the same IND grouping are enriched for co-occurrence in the same SE grouping (OR=212.37). We present two examples to illustrate the kind of insights this network enables: identification of drugs with shared IND but distinct SE genes as repurposing candidates, and identification of drugs with shared SE but distinct IND genes to assist treatment selection in patients with comorbidities.

Finally, we develop a neural network that directly links drug molecular effects onto disease genes and learns a gene-level score that quantifies each gene’s relative contribution to drug therapeutic versus side effects on disease.

## Introduction

Approximately 90% of clinical trials fail with efficacy and safety concerns accounting for the majority of attrition^1–5^. Efficacy failures often reflect suboptimal target selection while side effects usually arise either from on-target drug activity in unintended tissues or off-target interactions with secondary pathways^1,6^. Together, these two failure modes point to a common need: better understanding of how drugs interact with human biology.

Human genetics is well-positioned to address this need. Genome-wide association studies have identified thousands of disease-associated variants, and drugs that target the corresponding genes are more likely to be therapeutically effective^7–9^ - and also more likely to cause the disease as a side effect^10^. However, because the same genetic signals underlie both drug outcomes, there is a need for methods that can disentangle which disease genes are more likely to mediate therapeutic benefit versus side effects. Such insights have the potential to advance drug discovery and personalized medicine.

In a previous work, we developed DraphNet, a framework that maps drug molecular effects onto genetically dysregulated disease genes, enabling mechanistic interpretation of known drug-disease indications or side effects, as well as prediction of new ones^11^. A key feature of this framework is that all predictions of drug indications or side effects are based on the inferred effects of a drug on disease genes, allowing the construction of two gene sets for each drug: one including genes mediating its therapeutic effects and another including genes mediating its side effects. Here, propose that this approach can not only explain the shared basis of drug indications and side effects, it can also help disentangle a drug’s therapeutic effects from its side effects. Supporting this hypothesis, we previously showed that genetic overlap between a drug’s indications and a disease can predict whether that disease is caused as a side effect by the drug or not, and vice versa^12^.

Here, we first systematically characterize the shared genetic basis between drug indications and side effects. We then leverage this shared architecture to construct a genetics-driven classification of drugs sharing a common genetic basis for their therapeutic indications, or for their adverse side effects. We present two examples to illustrate the power of this approach in informing drug repurposing and personalized medicine. Finally, we develop a neural network model that jointly learns the genetic mechanisms for drug indications and side effects, designed to capture both shared genetics and the genes distinguishing beneficial from harmful effects.

## Results

### Supporting a shared genetic basis between drug indications and side effects

We find that drug indications and side effects are statistically associated, meaning a drug that treats a disease is more likely to also cause that disease as a side effect (odds ratio=2.52, p=2.28e-41). First, we ask whether Draphnet^11^ is able to model the shared genetic basis of both types of drug effects. To this end, we train two Draphnet models: one trained to predict drug indications and one predicting side effects. Briefly, each model integrates *in vitro* assayed molecular effects of 327 drugs (EPA ToxCast data^13,14^) with genetically dysregulated genes for 177 diseases (PhenomeXcan data^15^) and data on known drug indications or side effects. The model maps each drug’s ToxCast profile to a profile of its inferred effects disease genes to learn how drugs affect disease.

We then apply each model to the task it was not trained on. However, some drug-disease pairs are annotated as both an indication and side effect (n=366 pairs), which could artificially inflate cross-model predictions. To address this, we remove these overlapping pairs, re-fit both models, and repeat the analysis. The indication model no longer predicts side effects (AUROC=0.496). In contrast, the side effect model retains significant predictive performance for indications (AUROC=0.556; p_permutation_<0.001) (**Figure 1A**). As side effects can arise when a drug’s on-target effects adversely impact another disease, they may provide more insight into indications than the reverse.

**Figure 1.**
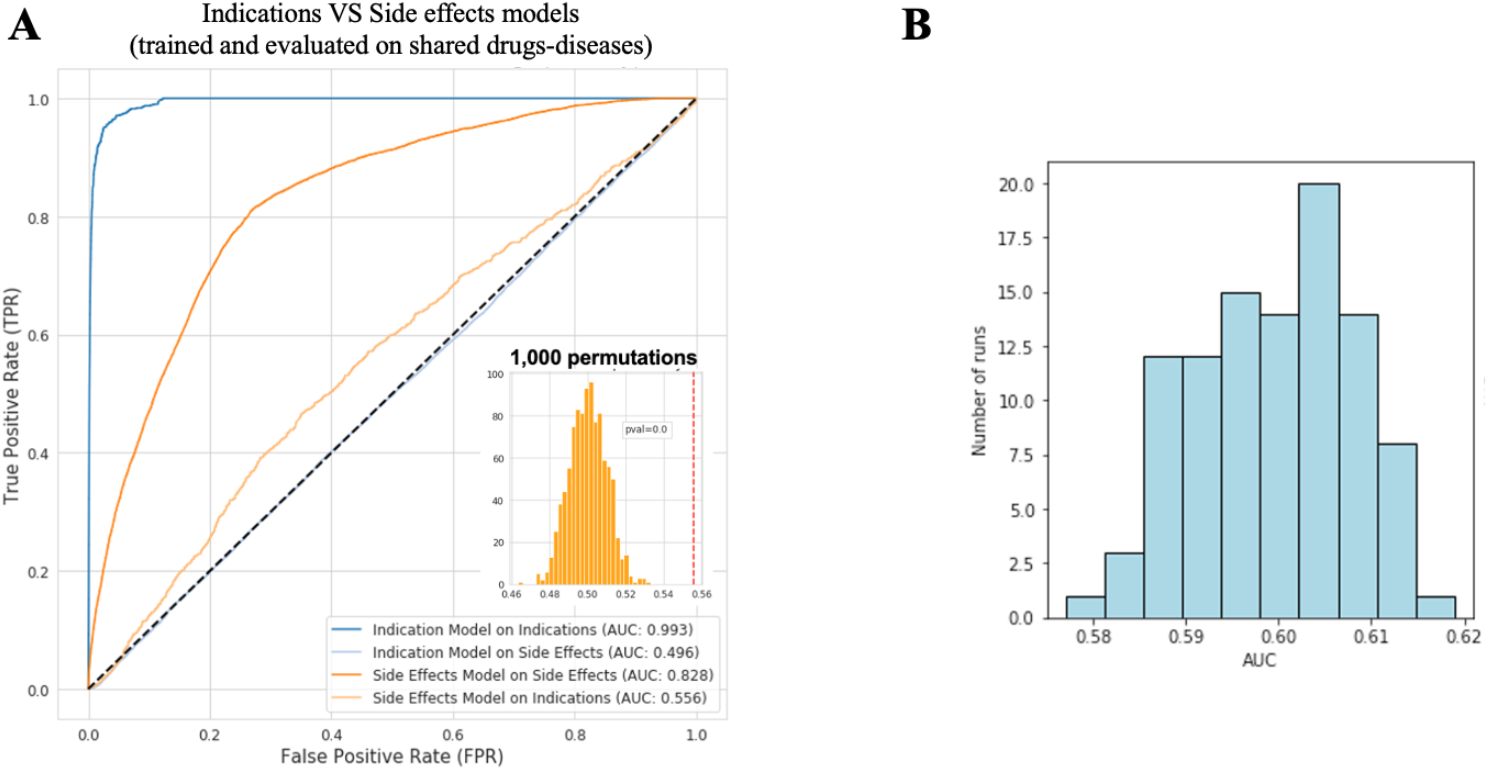
Cross-model prediction reveals a shared genetic signal between drug side effects and indications. **A**. Receiver operating characteristic (ROC) curves for cross-model prediction after removing drug-disease pairs annotated as both an indication and a side effect (n=366 pairs). Blue curves show the indication model predicting indications (dark blue) or side effects (light blue). Orange curves show the side effect model predicting side effects (dark orange) or indications (light orange). The inset histogram shows the null distribution of AUROCs obtained by randomly shuffling true drug indication labels (1,000 permutations) when using the side effect model to predict indications. The red dashed vertical line indicates the observed AUROC. **B**. Distribution of AUROCs from 100 iterations in which 8,020 positive side effect training labels are randomly removed to match the number of positive indication labels. All AUROCs exceed 0.5, indicating that the cross-prediction signal is not an artifact of class imbalance.

One alternative explanation for the stronger performance of the side effect model is that it has access to more positive training examples than the indication model (9,177 versus 1,157 true drug-disease labels). To exclude this possibility, we randomly remove 8,020 positive training examples from the side effects dataset to match the number of positive examples in the indication dataset, re-fit the model, and evaluate its ability to predict indications. We repeat this process 100 times and find that every subsampled model still predicts indications better than chance (AUROC>0.5) (**Figure 1B**), implying that the cross-prediction signal is not an artifact of class imbalance. Because our model relies on mapping each drug to the set of disease genes it impacts, these results suggest that drug indications and side effects have a common genetic basis.

### Shared genes mediate drug therapeutic and side effects

Next, we investigate the sets of genes our model infers to underlie each drug’s beneficial effects (IND genes) and its harmful effects (SE genes). We ask: do the same genes explain both types of effect? To test this, for each drug with at least one associated IND and SE gene (n=285), we quantify the overlap between its IND and SE genes. We compare this quantity to the overlap between that drug’s IND genes and the SE genes of every other drug. For more than half of the drugs (54.7%; 156/285), their IND and SE genes are more similar to each other than to those of other drugs (**Figure 2A**). In order to show that this is not just due to the fact that the model uses the same input ToxCast drug profile to infer a drug’s IND and SE genes, we train a set of null Draphnets on scrambled drug side effect labels. We use each null model to project the input ToxCast profile to the space of predicted side effect genes for a drug, creating a null SE gene set. Then, we can ask if the IND and SE genes learned by the model overlap more than this distribution of null SE gene sets. For 76% of the drugs (231/304), the observed overlap significantly exceeds the null (p_permutation_<0.05) (**Figure 2B**), suggesting that the within drug IND-SE gene overlap is due to the model learning the shared genetic basis of a drug’s indications and side effects.

**Figure 2.**
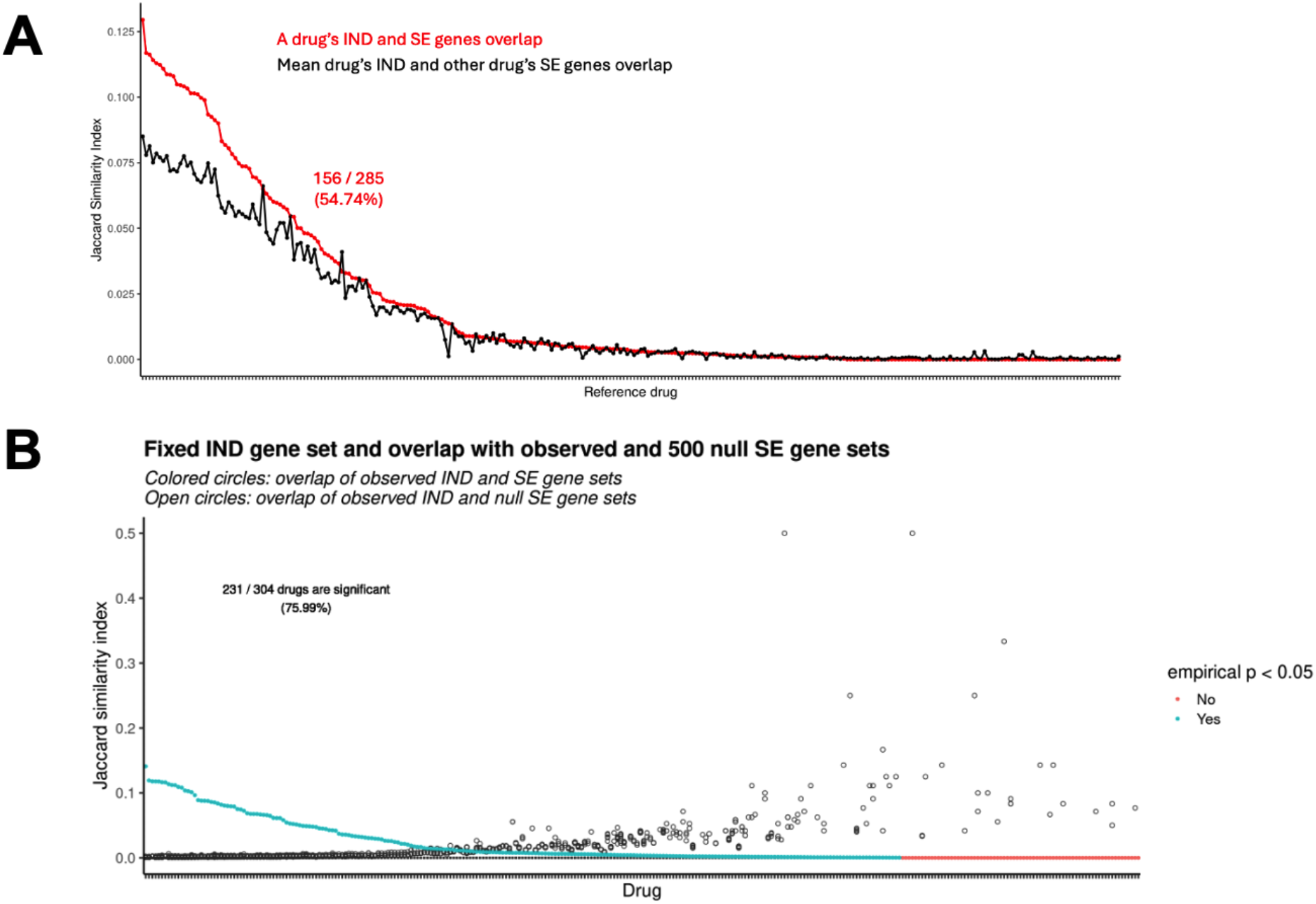
Genes mediating a drug’s therapeutic effects significantly overlap with those mediating its side effects. **A**. Within-drug versus cross-drug IND–SE gene overlap across all drugs with at least one associated gene (n = 285). For each drug (x-axis), the red line shows the overlap (y-axis) between its IND and SE gene sets (within-drug overlap), and the black line shows the average overlap between its IND genes and the SE genes of all other drugs (cross-drug overlap; used as baseline). Drugs for which the red line exceeds the black line have more similar IND and SE genes than expected from cross-drug comparisons. **B**. Comparison of observed versus null IND-SE gene overlap for all drugs included in the null model analysis (n=304). For each drug (x-axis), the solid point shows the observed overlap between its IND and SE gene sets (y-xis), and the open circles show the distribution of overlaps between its observed IND gene set and 500 null SE gene sets (generated by shuffling true drug-disease side effect labels). Solid points are colored blue if the observed overlap is significantly greater than the null distribution (ppermutation<0.05) and red otherwise.

### Drugs with more similar indication disease genes have more similar side effect disease genes

We next hypothesize that drugs with more similar therapeutic effects on disease genes also have more similar side effects on disease genes. To test this, we calculate pairwise similarities between all drug pairs based on (i) their IND genes and (ii) their SE genes. We observe a strong, positive correlation (Spearman ρ= 0.57; p<1e-300) between these two similarity measures across all drug pairs. Again, to ensure that this observation is not driven by patterns that simply exist in the input data, we compute pairwise drug-drug similarities derived from 500 null IND and SE gene sets. We find that the observed correlation significantly exceeds the null (**Figure 3**), suggesting that drugs with more similar indication biology also have more similar side effect biology.

**Figure 3.**
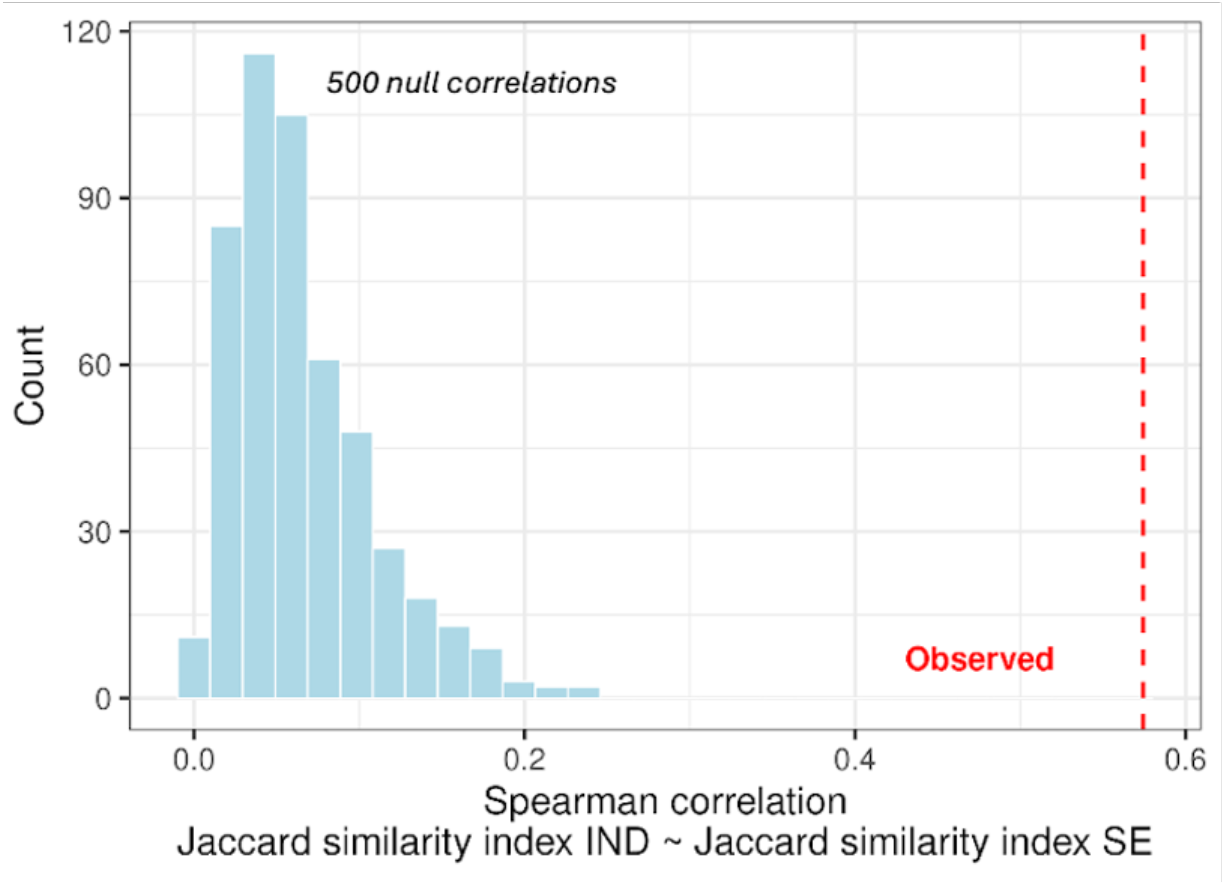
Drugs with similar indication gene profiles also have similar side effect gene profiles. The red dashed line indicates the observed correlation between the IND gene-based and SE gene-based overlap among drug pairs. The histogram shows the null distribution of correlations obtained from 500 null IND and SE gene sets, in which true drug-disease labels were shuffled prior to model training. The observed correlation significantly exceeds the null distribution.

### Drug classification based on shared effects on disease genes produces biologically meaningful cliques

The shared genetic architecture identified above raises a practical question: can we identify groups of drugs where indication and side effect genetics are aligned, and, more interestingly, groups where they diverge. Drugs sharing IND effects but belonging to distinct SE groups, represent repurposing candidates with different safety profiles. On the other hand, drugs sharing the same SE effects but belonging to distinct IND groups can inform treatment selection in patients with comorbidities by reducing toxicity risk. To investigate these drug groupings, we filter for genes associated with fewer than 15 drugs in order to use information from the most informative genes. Additionally, we only keep drugs with at least one IND and one SE gene. These result in a final set of 255 drugs associated with 2,691 IND genes and 2,467 SE genes, of which 630 are shared between the two sets. Using these gene sets, we construct a drug-drug network by connecting drug pairs with significant overlap in their IND genes (see Methods for details). We then identify fully connected cliques of drugs (minimum size of 3), referred to as IND cliques. We repeat this procedure using drug SE gene sets to create SE cliques of drugs. We find that drug triplets belonging to the same IND clique are significantly more likely to also co-occur in the same SE clique (OR=197.16; 95% CI: 172.94 - 225.15; Fisher’s exact test, two-sided). To ensure that this association is not driven by the 630 genes shared between IND and SE, we repeat the analysis after excluding these genes: the enrichment remains (OR=212.37; 95% CI: 183.56 - 247.79; Fisher’s exact test, two-sided).

Besides cliques of drugs that share both IND and SE genes, our classification framework reveals drugs with distinct beneficial and adverse effect mechanisms. We present two examples as a simple application of our approach:

#### IND clique example - drug repurposing

Dipyridamole, clofarabine and bosentan co-occur in the same IND clique but do not share a SE clique (**Figure 4A**). Clofarabine is used to treat acute lymphoblastic leukemia^16^, whereas dipyridamole and bosentan act on the cardiovascular system as an anti-platelet for stroke prevention^17^ and an endothelin antagonist for pulmonary hypertension^18^, respectively. Although this IND clique appears counterintuitive, examination of the IND genes shared by at least two members of the clique reveals a convergent biological pathway. Among the shared genes are *ACSS2*, which drives senescence-associated secretory phenotype by limiting purine biosynthesis^19^, and *G6PC3* and *RPIA*, both of which contribute indirectly to nucleotide synthesis via the Pentose Phosphate Pathway^20–23^. Together, these observations suggest that dipyridamole and bosentan may have anti-cancer effects by modulating genes involved in de novo nucleotide synthesis. Interestingly, this is the mechanism of action of clofarabine^24^, the third drug in this IND clique. Supporting this repurposing hypothesis, prior studies link dipyridamole to a reduced risk of lymphoid neoplasms^25^ and report anti-cancer effects of bosentan^26^.

**Figure 4.**
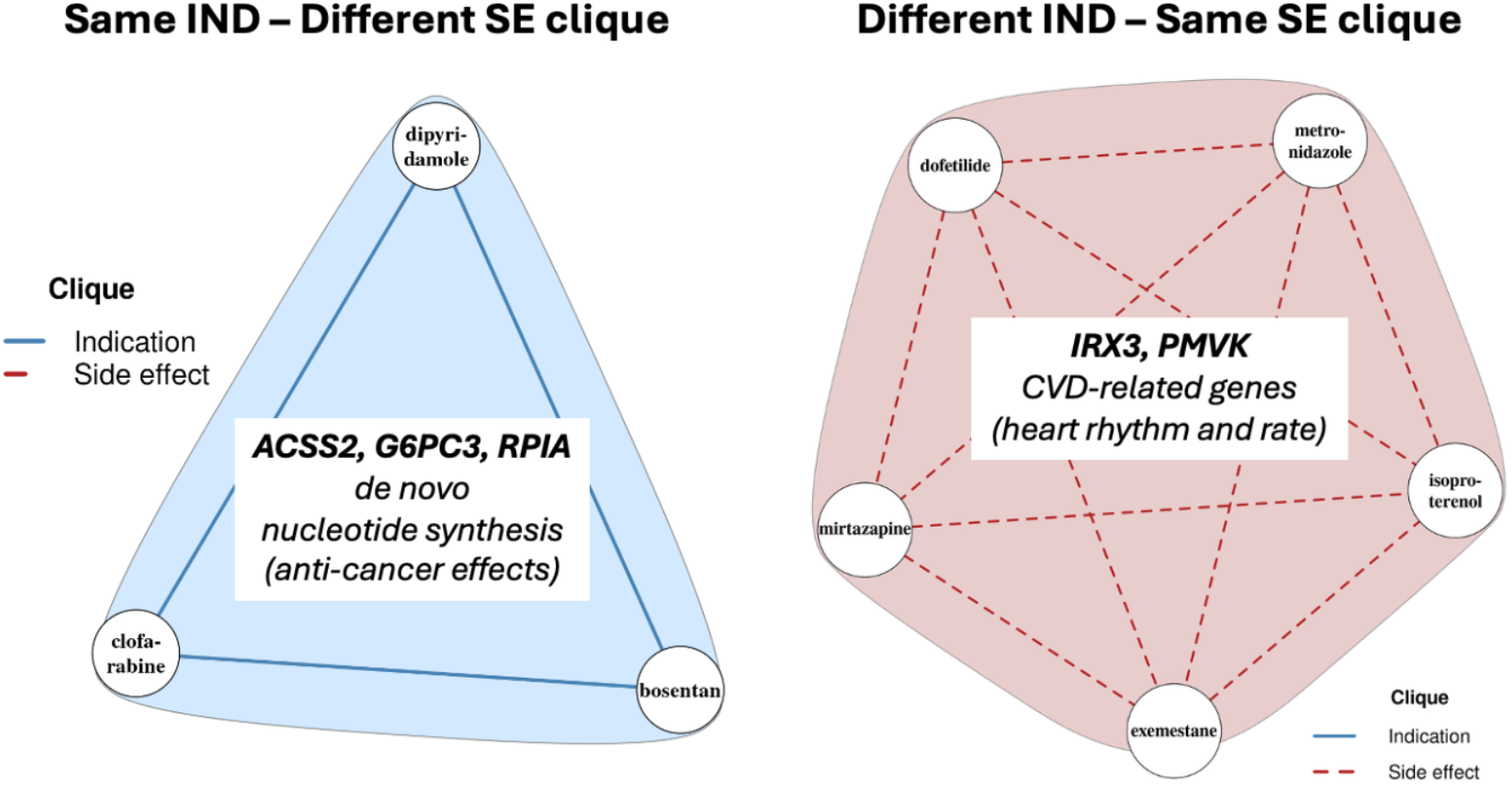
Genetics-based drug cliques reveal shared therapeutic and adverse effect biology, supporting drug repurposing and toxicity-aware prescribing. **A**. Example of drugs sharing an IND clique (blue) but belonging to different SE cliques. Dipyridamole (antiplatelet, stroke prevention), bosentan (endothelin antagonist, pulmonary hypertension), and clofarabine (purine nucleoside antimetabolite, acute lymphoblastic leukemia) share IND genes including *ACSS2* (purine biosynthesis), *G6PC3*, and *PRIA* (Pentose Phosphate Pathway), suggesting convergence on a nucleotide synthesis pathway. **C**. Example of drugs sharing an SE clique (red) but belonging to different IND cliques. Dofetilide, exemestane, isoproterenol, metronidazole, and mirtazapine share a cardiovascular adverse effect profile despite distinct therapeutic indications spanning infectious disease, oncology, psychiatry, and cardiology. Shared SE genes *IRX3* and *PMVK*, associated with hypertension, atrial fibrillation, and coronary artery disease, are identified as signature genes of this clique.

#### SE clique example - personalized medicine

We identify a SE clique including dofetilide, exemestane, isoproterenol, metronidazole and mirtazapine (**Figure 4B**). All five drugs are known to induce cardiac risk by affecting the heart rate and rhythm, which likely explains their SE clustering. *IRX3* and *PMVK*, genes associated with hypertension, atrial fibrillation and coronary artery disease, all cardiovascular diseases (PhenomeXcan p_BH_adjusted_<0.1), are found to be affected by most of the drugs in the clique. This supports the biological relevance of this SE clique while providing specific genes that may mediate these drug side effects. Despite this shared side effect profile, these drugs have different therapeutic uses: metronidazole is used for bacterial infections, mirtazapine is an anti-depressant and exemestane is an aromatase inhibitor used in breast cancer treatment. Dofetilide and isoproterenol are both used to treat abnormal heart rates, but in opposite directions: dofetilide slows nerve impulses in the heart, whereas isoproterenol treats bradycardia by increasing heart rate.

### Joint modeling of drug therapeutic and side effects reveals outcome-specific disease genes

Although IND and SE gene sets for a drug significantly overlap, we also identify genes specific to efficacy or toxicity. Distinguishing these gene sets is important as it can guide the prioritization of safer therapeutic targets during drug development. Here, we develop a neural network model that uses a shared drug representation to jointly predict indications and side effects. The model learns three different sets of parameters to make drug-disease predictions: a shared component (W_SHARED_) which captures information relevant to overall drug effects, and two-task specific components, W_IND_ and W_SE_, which capture signals associated with indications and side effects, respectively (see Methodology for details about model architecture and training; **Figure 6**).

We first evaluate how well each component contributes to predictions. For held-out drug-disease indications, the model performs well when combining W_IND_ + W_SHARED_ (AUROC_IND_ = 0.78) and also shows strong performance when each component is used individually (AUROC_IND, Wind_ = 0.75; AUROC_IND, Wshared_ =0.74) (**Figure 5A**). This suggests that both W_IND_ and W_SHARED_ are relevant for indication predictions. Additionally, the improvement observed when combining both components suggests that they capture complementary information. We observe a similar pattern for side effect prediction (AUROC_SE, Wse + Wshared_ = 0.85; AUROC_SE, Wse_ = 0.82; AUROC_SE, Wshared_ = 0.78; **Figure 5B**). As expected, W_SHARED_ is predictive across both tasks as it captures signals common to drug-disease indications and side effects.

**Figure 5.**
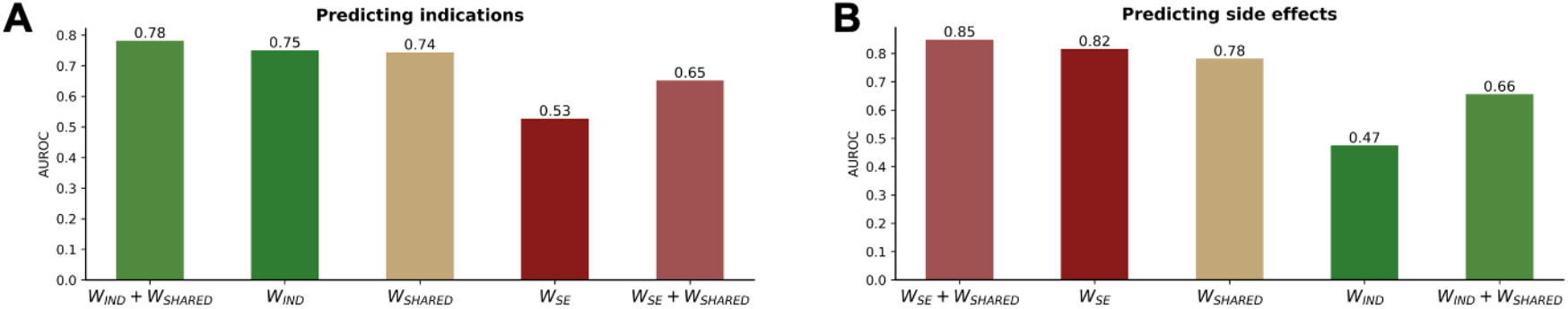
Our neural network model learns gene-specific scores that quantify how each disease gene contributes to drug therapeutic and adverse effects and directly captures the common genetic basis between both drug outcomes. Performance of indication-specific (W_IND_), side effect specific (W_SE_) and shared (W_SHARED_) gene scores in predicting drug indications (**A**) and side effects (**B**). The shared component W_SHARED_ predicts both drug outcomes while the task-specific components W_IND_ and WSE specialize in the outcome they were trained on.

We next ask whether W_IND_ and W_SE_ are specific to their respective tasks. If so, we expect each component alone to perform poorly when applied to the task it was not trained on. Consistent with this, using W_IND_ alone to predict side effects results in near random performance (AUROC_SE, Wind_ = 0.47). However, adding W_SHARED_ restores predictive power (AUROC_SE, Wind + Wshared_ = 0.66) but it remains lower compared to using W_SE_+ W_SHARED_ (**Figure 5B**), indicating that W_IND_ contains indication-specific signals not relevant for side effect prediction. The same pattern is observed when using W_SE_ or W_SE_ + W_SHARED_ to predict indications (**Figure 5A**).

Together, these results demonstrate that our model effectively disentangles shared and task-specific genetic signals. Importantly, since these W matrices are diagonal and match the dimensionality of the disease gene space (10,027 genes), the learned weights can be interpreted as gene-level scores quantifying each gene’s contribution to a drug’s therapeutic and adverse effects on disease.

## Discussion

Previous studies have shown that human genetics is predictive of both drug indications^7–9^ and side effects^27^, suggesting a shared biological basis. However, this overlap has not been systematically quantified or leveraged for target prioritization. Here, our analysis across 327 drugs and 177 disease phenotypes provides evidence that therapeutic and adverse drug outcomes share a common genetic architecture.

While this shared basis is informative, it also presents a key challenge: drug development requires distinguishing between beneficial and harmful effects to prioritize targets with favorable risk-benefit profiles and reduce clinical attrition. To address this, we developed a neural network model that quantifies the contributions of disease-associated genes to both drug indications and side effects. Our model captures shared genetic signals while simultaneously identifying outcome-specific components, suggesting that certain genes are more likely to mediate therapeutic effects, whereas others are more strongly associated with toxicity. Importantly, our model learns interpretable gene-level scores, providing a practical tool for target prioritization in drug discovery.

Current drug classification systems, such as the Anatomical Therapeutic Chemical (ATC) system, group drugs based on organ systems they act upon, therapeutic intent, pharmacological properties, and chemical structure^28^. While useful, these systems do not capture the full spectrum of drug-induced phenotypic consequences as they largely overlook drug side effects^28–31^. Here, we introduce a genetics-driven drug classification based on shared gene-level mechanisms underlying adverse effects. This classification enables grouping drugs by their molecular impact on disease biology, rather than their intended use. Such a classification has potential applications in personalized medicine, particularly for patients with comorbidities. For instance, we identify a cluster of drugs including ofetilide, exemestane, isoproterenol, metronidazole and mirtazapine. Although these drugs are prescribed for different indications, they all cause cardiovascular side effects. This means that co-administration of these drugs may increase the risk of cardiovascular toxicity. This example illustrates how genetics-based drug classification can inform safer prescribing decisions and support more personalized treatment strategies

Our work has several limitations. First, we are unable to distinguish on-target from off-target side effects due to the lack of comprehensive annotations. Second, although the neural network yields gene-level scores, their interpretation is not entirely straightforward, as they can take both positive and negative values. Future work could improve interpretability by constraining these scores (e.g., enforcing non-negativity or bounding them within [0,1]) or by exploring alternative model architectures. Nonetheless, our framework provides a foundation for disentangling the genetic contributions to therapeutic and adverse drug effects. Third, the datasets used introduce biases. ToxCast captures i*n vitro* drug bioactivity and does not fully reflect organ- or organism-level effects, potentially limiting the biological fidelity of the learned representations.

Similarly, PhenomeXcan relies on GWAS data to infer gene-disease associations, but GWAS do not capture the full spectrum of genetic contributions to disease, leading to incomplete gene coverage^32^. Incorporating other comprehensive resources, such as DisGeNET^33^, could help address this limitation in future work. Finally, the SIDER database was last updated in 2015 and is known to be incomplete and affected by reporting biases, which influence what our model learns. Although it was the only readily accessible source of side effect labels at the time, future studies could leverage more recent resources, such as onSIDES^34^.

In conclusion, we demonstrate that drug indications and side effects are not independent processes but instead share a common genetic basis. We further introduce a genetics-driven drug classification that groups drugs according to shared side-effect biology, addressing a key limitation of existing systems that largely overlook adverse effect profiles. In addition, we develop an interpretable neural network that jointly models drug-disease relationships while explicitly disentangling shared and outcome-specific gene contributions, providing a principled framework for understanding drug action at the gene level. Together, these findings underscore the value of integrating therapeutic and toxicity signals within a unified framework of drug and disease biology, with the potential to guide safer and more effective drug discovery.

## Methods

### Data preparation and implementation of affinity regression for binary outcomes

We have previously described the preprocessing of EPA ToxCast data and the disease genetic gene expression profiles. We also implement affinity regression for binary outcomes as outlined in our prior work and construct drug gene sets by mapping drug effects onto the disease genome. Finally, we apply our previously described framework to cluster drugs based on their indication and side effect gene sets (clique analysis)^11^.

### Model extension using neural network to predict both drug outcomes and learn gene-level scores

Our goal is to learn gene-level scores that quantify each gene’s contribution to the therapeutic and adverse effects of drugs on disease, while explicitly capturing shared contributions between these outcomes. To achieve this, we develop a neural network model that projects drug molecular profiles from ToxCast *D* matrix into a gene-level latent space *L*. Then, it uses that projection to link drugs onto disease genes, such that if a drug impacts a gene that is strongly linked to a disease, then the drug is predicted to impact that disease.

The model uses an overcomplete autoencoder to project the *D* into *L*. The encoder consists of fully connected layers with ReLU activations and dropout regularization and maps each drug to a 10,027 dimensional latent representation, which matches the number of disease genes in the *P* matrix. A symmetric decoder reconstructs the input *D* from *L*. The model is optimized using the mean square error (MSE) loss between the input data and the reconstructed output. As mentioned earlier, the *D* matrix has missing values. Although we generated a complete *D* matrix using SoftImpute, imputed values may include noise. To avoid learning that noise, we compute the reconstruction MSE loss only over originally observed values in *D*.

To model drug-disease relationships, the latent representation *L* is linearly mapped to disease outcomes using a logistic regression-like layer. Specifically, predictions are generated as LWP^t^, where W captures gene-level contributions linking drug effects to disease genes. To disentangle shared and task-specific effects, the model learns three diagonal weight matrices: a shared component (W_SHARED_), an indication-specific component (W_IND_) and a side effect-specific component (W_SE_). Indications are predicted using *L*(*W*_*SHARED*_*+W*_*SE*_)*P*^*t*^ while side effects are predicted using *L*(*W*_*SHARED*_*+W*_*IND*_)*P*^*t*^. The diagonal structure of learned Ws ensures that each parameter directly corresponds to a disease gene, enabling interpretable gene-level scoring of therapeutic and adverse effects. We prompt sparsity in W_IND_ and W_SE_ by applying L1 regularization. We do not L1 regularize the W_SHARED_ to allow capturing of common signals without constraints. **Figure 6** shows the full model design. Below, we mention details about hyperparameter tuning and model training.

**Figure 6.**
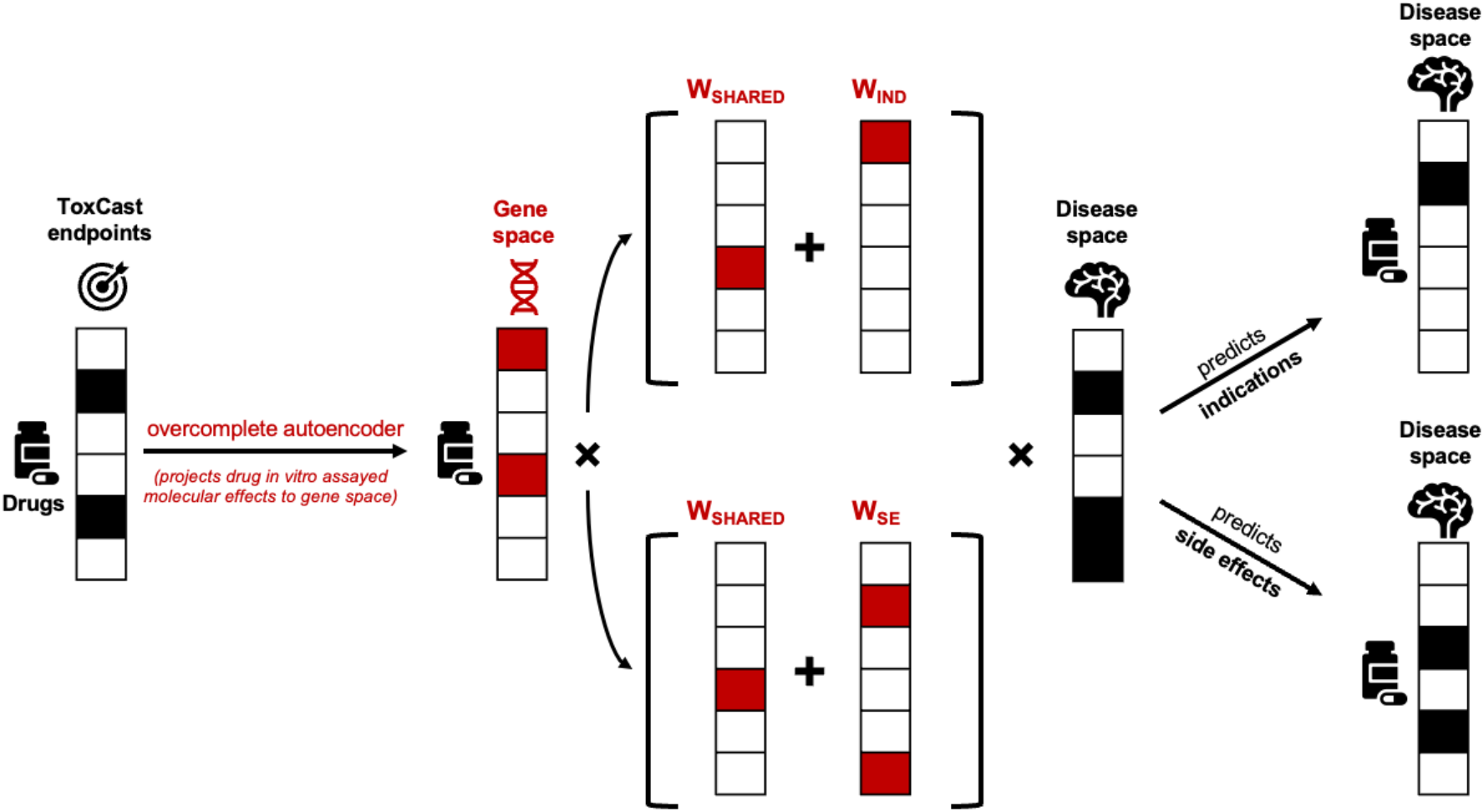
Neural network model architecture. Red color indicates what the model learns. Black color indicates data used as input for training.

The model is trained jointly using three loss functions: an MSE loss for the autoencoder and two binary cross-entropy losses (BCEWithLogitLoss) for predicting indications and side effects. The encoder is updated by all three losses, the decoder only by the reconstruction MSE loss, the W_SHARED_ by both classification losses, and W_IND_ and W_SE_ by their respective task-specific losses. To balance these objectives and prevent any single loss from dominating the learning process, we weight each loss term and treat those weights as hyperparameters.

Model hyperparameters include the learning rate, architecture of the autoencoder (number and size of hidden layers, dropout rate), L1 regularization strength (λ_IND_, λ_SE_), and loss weights. To tune these, we split the drugs into training (80%), validation (10%) and test (10%) sets. We train models across different hyperparameter combinations on the training set and select the optimal configuration based on the highest combined performance on the validation set (AUPRC_IND_ + AUPRC_SE_). To prevent information leakage arising from drugs with similar ToxCast profiles appearing in different splits, we construct split the data using a clustering-based approach. Specifically, we binarize the ToxCast D matrix (values ≥ 0.8, activity threshold recommended by ToxCast) and compute pairwise Jaccard distances between drugs. Using this distance matrix, we applied k-medoids clustering to group drugs with similar ToxCast profiles, selecting the optimal number of clusters based on silhouette score (k=73). We further merge the resulting clustered into eight approximately balanced groups using a greedy procedure that preserves cluster structure while equalizing group sizes. Finally, we assign entire groups to the training, validation and test sets to achieve a 80:10:10 split while ensuring that similar drugs remain within the same split.

The final model is trained on the combined training and validation set using Adam optimizer and class weights to address class imbalance in the Y_IND_ and Y_SE_ (parameter pos_weight = Y_positive_ / Y_negative_). Its generalizability is evaluated on the test set. The selected learning rate is 0.0001. The autoencoder architecture consists of a single hidden layer with 4,096 units and a dropout rate of 0.2. L1 regularization parameters are λ_IND_=λ_SE_=0.0001 and loss weights are w_MSE_=0.8, w_BCE, IND_=0.1, w_BCE, SE_=0.1.

## Data Availability

All data produced in the present study are available upon reasonable request to the authors and will be made available in this GitHub repository: https://github.com/lalagkaspn

